# Home-Based Pulsed Electromagnetic Field Therapy for Blood Pressure and Cardiometabolic Health: A Systematic Review and Meta-Analysis

**DOI:** 10.64898/2026.09.23.26363775

**Authors:** Reuben R. Simfukwe, Pearl Aovare, Felix P. Chilunga

## Abstract

**Background:** Cardiometabolic disease is a leading cause of death, and most therapies suppress established pathology rather than restore functional capacity. Pulsed electromagnetic field (PEMF) therapy is a non-invasive stimulus touted as a potential complementary approach that may engage adaptive responses, with practical potential for home-based use. We systematically reviewed and meta-analysed PEMF effects on cardiometabolic outcomes.

**Methods:** We searched seven electronic databases and Google Scholar from inception to 31 August 2026 for studies of PEMF in adults, healthy or with cardiometabolic conditions. Between-group differences were rescaled into minimal clinically important difference (MCID) units to facilitate comparison across diverse outcomes. Established anchor-based MCIDs were used where available; otherwise, 0.5 pooled within-arm SD was used. Effects were pooled using multilevel random-effects models with study-clustered robust variance, with subgroup, sensitivity, and exploratory dose-response analyses. The review was registered with PROSPERO (CRD420261404575).

**Results:** Ten studies (353 participants) were included. PEMF was associated with an overall benefit of 0.54 MCID units (95% CI 0.08–1.00). Effects were largest in hypertension (1.24 MCID units, 0.52–1.96), with no observed heterogeneity across two double-blind trials (I^2^=0%). Exploratory multivariable meta-regression associated greater cumulative exposure and lower frequency with larger effects.

**Conclusions:** PEMF therapy may offer a non-invasive, non-pharmacological adjunct for improving cardiometabolic outcomes, with potential for home-based use. The most compelling signal was observed in hypertension, where the pooled effect exceeded one clinically important difference, although confirmation in larger trials is required.

## INTRODUCTION

Cardiometabolic diseases, which include hypertension, type 2 diabetes, obesity, and cardiovascular disease, are among the leading causes of death worldwide, and their burden continues to rise despite decades of investment in prevention and treatment^1^. Although these conditions carry distinct clinical labels, they share a common pathophysiological thread: a progressive erosion of the body’s capacity to adapt to physiological stress and maintain homeostasis. This manifests as impaired glycaemic regulation, endothelial dysfunction and arterial stiffening, chronic low-grade inflammation, and metabolic inflexibility^2^. As this adaptive reserve declines, disease progresses and becomes increasingly difficult to reverse.

Pharmacotherapy remains the mainstay of management, yet most agents act by continuously suppressing a pathological process rather than restoring the body’s own regulatory function^3^. Sustained exposure can lead to tolerance, adverse effects, and polypharmacy, all of which undermine long-term adherence^3,4^. These limitations are felt most acutely in hypertension, the single largest modifiable contributor to cardiovascular risk^5^, where a substantial proportion of patients remain above target despite multi-drug regimens^6^. There is therefore growing interest in safe, non-pharmacological interventions, particularly self-administered ones (home-based), that could complement medication (adjunct non-invasive therapies)^3^.

Pulsed electromagnetic field (PEMF) therapy is one such intervention. A PEMF device is a home-based device that consists of a coil or planar applicator, often incorporated into a mat, wrap, or handheld unit, that is positioned on or near the body and driven by brief pulses of electric current. Each pulse generates a rapidly changing magnetic field that penetrates tissue largely unattenuated by skin, adipose tissue, or bone and, through electromagnetic induction, induces low-amplitude electrical currents within the tissue^7,8^. These induced currents are thought to modulate cellular activity, including transmembrane ion transport^9,10^, nitric-oxide signalling^9^, inflammatory pathways^11^, and microvascular perfusion^12^, the same processes disrupted in cardiometabolic disease^2^. The fields are of low intensity (typically microtesla to millitesla) and low frequency (a few to a few hundred hertz), and are delivered in short sessions lasting minutes^7,8^.

Two features make PEMF attractive as a cardiometabolic adjunct. First, it is non-invasive, pharmacologically inert, and amenable to self-administration (home-based), which may mitigate the adherence barriers that limit drug therapy^8,10,13^; it also carries an established safety record in other indications, notably fracture healing^14^. Second, because the stimulus ceases the moment the device is switched off, PEMF is inherently intermittent, delivering repeated cycles of transient stress followed by recovery. This exposure-recovery pattern is characteristic of hormesis, a biphasic response in which mild, transient stress elicits adaptive strengthening rather than injury^10,15^, offering a plausible route to restoring the adaptive capacity that cardiometabolic disease erodes. Its precise mechanisms, however, remain incompletely defined^7^.

Despite this promise (biological effects, home-based use, non-invasive nature), the human evidence for PEMF in cardiometabolic disease has never been collated to adequately quantify its effectiveness. To address this gap, we estimated the effects of PEMF on cardiometabolic outcomes (hypertension, type 2 diabetes, obesity, and cardiovascular disease) overall and within subgroups.

## METHODS

### Study design and registration

We conducted this systematic review and meta-analysis in accordance with the Preferred Reporting Items for Systematic Reviews and Meta-Analyses (PRISMA) 2020 statement^16^ and registered the protocol prospectively in the International Prospective Register of Systematic Reviews (PROSPERO; CRD420261404575). The review evaluated whether PEMF therapy is associated with changes in cardiometabolic outcomes in adults aged 18 years or older.

### Eligibility criteria

Eligible studies enrolled adults (≥18 years), either healthy or with a cardiometabolic condition (hypertension, diabetes, obesity, metabolic syndrome, or cardiovascular disease), evaluated PEMF therapy, and reported at least one post-exposure outcome among systolic or diastolic blood pressure, fasting glucose, glycated haemoglobin, insulin-resistance indices, body mass index (BMI), vascular function, arterial stiffness, or inflammatory markers. All study designs were considered, from randomised controlled trials to observational studies; animal and in vitro work and non-English publications were excluded.

### Information sources and search strategy

We searched seven electronic databases (PubMed, Embase, Web of Science, Scopus, the Cochrane Central Register of Controlled Trials, ClinicalTrials.gov, and the WHO International Clinical Trials Registry Platform) from inception to 31 August 2026, together with Google Scholar for grey literature. We also hand-searched the reference lists of included studies and relevant systematic reviews. No date restrictions were applied. Search terms combined controlled vocabulary (Medical Subject Headings and Emtree terms) with free-text terms for pulsed electromagnetic exposure and cardiometabolic outcomes; the full search strategy for each database is provided in the Supplementary Material (Table S1).

### Study selection and data extraction

Records were imported into Zotero for de-duplication, and a structured Microsoft Excel template was applied uniformly to every study. To support outcome-level meta-analysis, the unit of extraction was the study × outcome (× subgroup where applicable) rather than a single summary row per study. We captured three groups of fields: study-level descriptors (first author, year, country, design, sample size, numbers allocated to the PEMF and control arms, mean age, percentage female, comparator type, and adverse events); PEMF exposure parameters (intensity in the originally reported units, frequency, single-session duration, sessions per week, total intervention duration, and any cumulative exposure); and, for each outcome, its name and unit, the intervention- and control-arm end-of-intervention means, standard deviations, and p-values, and any reported between-group mean difference.

Two reviewers (F.P.C. and R.R.S.) independently screened the records and extracted the data from each published report and its supplementary material, recording every outcome as a separate row and repeating the study-level and exposure fields; disagreements were resolved by discussion, with a third reviewer (P.A.) adjudicating when consensus could not be reached. Where a study reported clinically distinct sub-populations (as when Sun et al. 2016^17^ and Wróbel et al. 2010^18^ reported PEMF effects separately for patients with diabetes and for healthy participants), each subgroup was extracted as a separate, labelled row so that pooling decisions remained transparent.

### Standardisation of PEMF exposure parameters

Because trials reported exposure in different units, we converted each variable to a common unit alongside its original value: intensity to microtesla (1 Gauss = 100 µT; 1 mT = 1000 µT), frequency to hertz (using the midpoint of any reported range, as for the 6 to 8 Hz protocol of Nishimura et al. 2011^19^), and single-session duration to minutes. Sessions per week were inferred from the reported schedule using conventional mappings (for example, “daily” as 7 and “five times per week” as 5), and total exposure was computed as single-session minutes multiplied by sessions per week and by weeks, with single-session studies (Sun et al. 2016^17^) assigned the one-session total. Both the original and standardised values were retained so that all conversions are independently auditable.

### Effect-size standardisation

The included trials measured outcomes on different scales (mmHg, mg/dL, arbitrary laser-Doppler units, millimetres, dimensionless ratios, etc.), so pooling raw mean differences was not possible. We therefore expressed every between-group mean difference in units of the minimal clinically important difference (MCID) for that outcome, following the approach by Norman, Sloan, and Wyrwich^20,21^. The MCID is the smallest change in an outcome that a patient or clinician would perceive as beneficial^21^; dividing an observed change by it places disparate measurements on one clinically interpretable scale, so that a value of 1.0 denotes exactly one clinically meaningful improvement and effects can be compared and combined across outcomes:

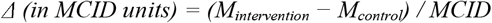

We preferred this MCID-based scaling to the more common standardised mean difference (Hedges’ g), which divides each effect by its own study’s standard deviation. Because that denominator varies with each sample’s heterogeneity, an identical true effect appears larger in a homogeneous trial and smaller in a more variable one, and the pooled estimate carries no natural clinical unit^21^. Dividing instead by a fixed, externally anchored MCID keeps the denominator constant across studies, so that a pooled value of 1.0 always denotes one clinically important difference, yielding an estimate that is at once comparable across disparate outcomes and directly interpretable at the bedside.

Where an established anchor-based MCID existed, we used it: 5 mmHg for systolic and diastolic blood pressure^22^. For fasting glucose (10 mg/dL), BMI (1 kg/m^2^), and the ankle-brachial index (0.05), for which no consensus anchor-based MCID has been established, we applied empirical thresholds chosen by the authors to represent the smallest change considered clinically meaningful. For the remaining outcomes without a recognised anchor (capillary diameter, microcirculation in arbitrary units, plasma nitric oxide, vein diameter, blood-flow velocity, cardiovascular-disease risk score, arterial-stiffness index, and defensin), we applied a distribution-based MCID of half the pooled within-arm standard deviation, in line with the widely used convention of 0.5 SD as a moderate effect^20^. The MCID source (anchor-based, empirical, or distribution-based) was recorded for every outcome.

All effects were direction-coded so that a positive value always denotes clinical benefit; for outcomes in which a decrease is desirable (blood pressure, fasting glucose, C-reactive protein, arterial-stiffness index, and cardiovascular-disease risk), we reversed the sign of the mean difference before standardisation. The standard error of each standardised effect followed from the independent-groups standard error of the raw mean difference, divided by the MCID. Two studies did not report the standard deviations required for this step (Wróbel et al. 2010^18^ for all outcomes; Biermann et al. 2020^23^ for most) and, although retained for the qualitative synthesis, contributed no data to the quantitative model. The use of MCIDs was not prespecified in the registered protocol and was introduced during the analysis to facilitate interpretation of the clinical relevance of treatment effects.

### Risk-of-bias assessment

Risk of bias was assessed at the study level with the Cochrane Risk of Bias 2 tool^24^ for randomised trials and ROBINS-I^25^ for the two non-randomised studies (Biermann et al. 2020^23^, a within-subject contralateral-control design; Ahmad et al. 2021^26^, a non-randomised allocation). We based each domain judgement on the published methods.

### Statistical analysis

For the primary analysis we fitted a multilevel random-effects meta-analysis, with outcomes nested within studies^27^, by restricted maximum likelihood using the metafor package in R (version 4.3.2)^28^. Because several trials contributed more than one outcome, we applied study-clustered robust (sandwich) variance estimation^29^ to account for the resulting within-study correlation, using the robust() function in metafor with its small-sample adjustment and t-distributed tests with degrees of freedom based on the number of studies. Each pooled effect is reported in MCID units with a 95% confidence interval (CI), a CI excluding zero indicating a statistically detectable effect and a value of 1.0 one full MCID; heterogeneity was summarised with the multilevel I^2^ statistic.

Two conditions had enough data for subgroup analysis: hypertension (five outcomes) and diabetes (ten outcomes); we tested for a between-subgroup difference by adding condition as a moderator and described single-study subgroups (e.g. metabolic syndrome; peripheral artery disease). Robustness was assessed in two ways: by restricting the analysis to double-blind, sham-controlled trials delivering a multi-session treatment course, and by omitting each study in turn (leave-one-out). A dose-response meta-regression examined PEMF intensity, frequency, single-session duration, total exposure, and intervention duration, each z-scaled and modelled singly, with intensity, total exposure, and frequency also modelled jointly; because the number of contributing studies was limited, we interpret its coefficients as candidate moderators to be confirmed rather than as definitive dose-response relationships^30^. Small-study effects were examined with Egger’s test^31^, which is underpowered with fewer than ten studies^32^, and the certainty of evidence was rated with the Grading of Recommendations Assessment, Development and Evaluation (GRADE) framework^33^.

## RESULTS

### Study selection and characteristics

We identified 66 records through academic databases, and one additional record through Google Scholar. After screening all 67 titles and abstracts, we excluded 47 records and assessed 20 full-text reports for eligibility, of which we excluded a further 10. Ten studies met all criteria and entered the systematic review^17–19,23,26,34–38^; two of these did not report standard deviations and so contributed to the qualitative synthesis but not the meta-analysis (Figure 1).

**Figure 1.**
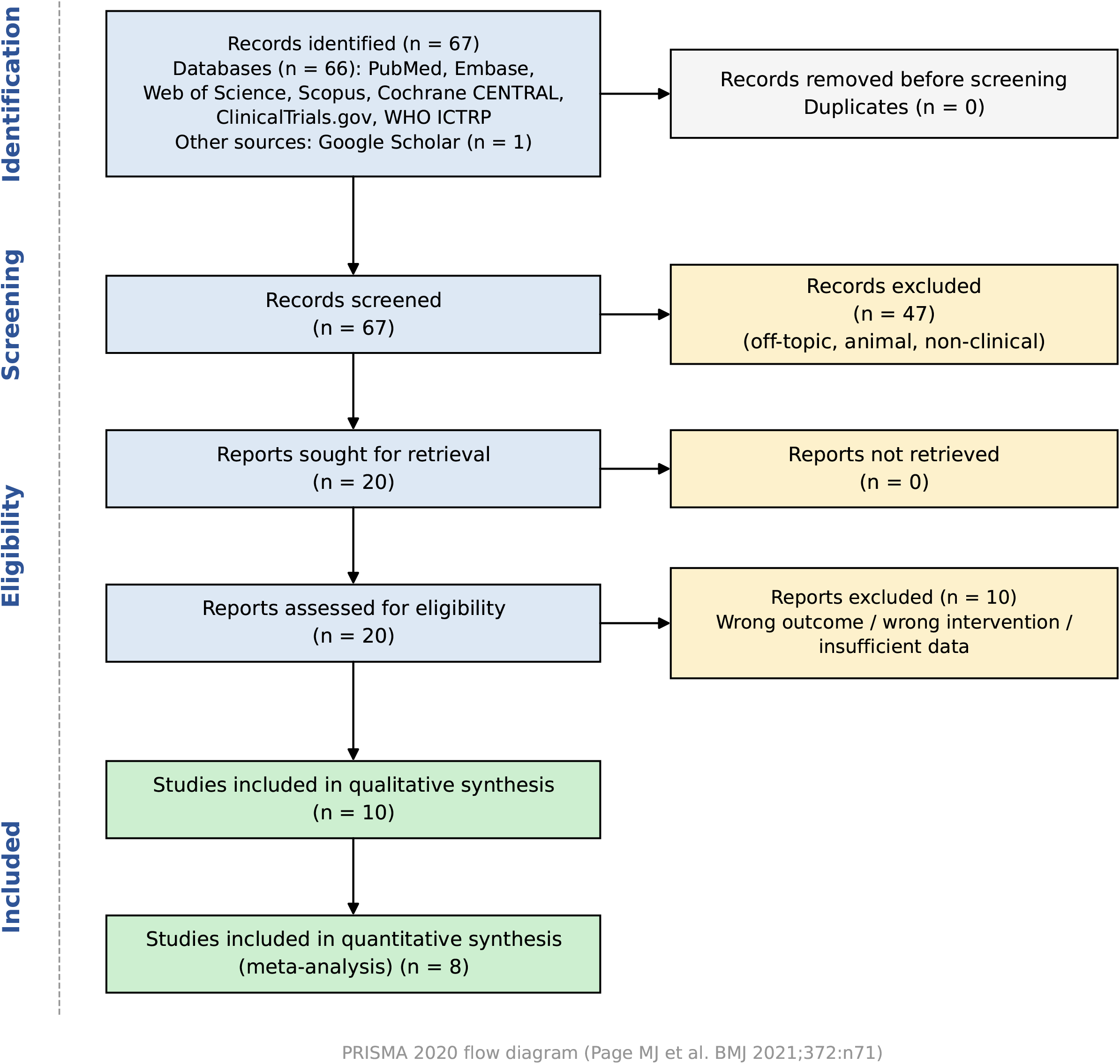
PRISMA 2020 flow diagram. Study identification, screening, eligibility assessment, and inclusion. Of 67 records identified (66 from databases and one from Google Scholar), 10 studies were included in the qualitative synthesis and 8 in the meta-analysis.

The ten included studies were published between 2010 and 2025 and together enrolled 353 participants, with mean ages ranging from 25 to 67.4 years. They were conducted across eight countries (Saudi Arabia, Poland, Hungary, the United States, China, Germany, Japan, and Egypt) and spanned healthy adults and patients with hypertension, diabetes, peripheral artery disease, and metabolic syndrome. Eight were randomised controlled trials and two were non-randomised. PEMF protocols varied widely in intensity (1 to 2000 µT), frequency (7 to 188.5 Hz), and session duration, with treatment courses ranging from a single exposure to 12 weeks. PEMF was well tolerated: only one study (Nishimura et al. 2011^19^) reported adverse events, mild paraesthesia of the hands in two participants receiving PEMF, which resolved spontaneously within days. Table 1 summarises the study characteristics, with further detail in Table S2.

**Table 1.** Characteristics of the 10 included studies.

| Study (year) | Country | Design | Population (n PEMF/control) | Age (y) | PEMF protocol (intensity; frequency; session; duration) | Key outcomes assessed |
| --- | --- | --- | --- | --- | --- | --- |
| Abdelaal et al. (2025)<sup>34</sup> | Saudi Arabia | RCT | Type 2 diabetes (14/14) | 57.8 | 2000 μT; 15 Hz; 30 min daily; 8 wk | Fasting glucose; cardiovascular-disease risk |
| Wróbel et al. (2010)<sup>18</sup> | Poland | RCT | Diabetic polyneuropathy (32/29) + healthy (20, PEMF only) | 49.8 | 100 μT; 188.5 Hz; 20 min ×5/wk; 3 wk | Defensin; CRP |
| Stewart et al. (2020)<sup>35</sup> | USA | RCT | Hypertension (15/11) | 59.5 | 390 μT; 30 Hz; 16 min thrice daily; 12 wk | Systolic/diastolic BP; nitric oxide |
| Rikk et al. (2013)<sup>36</sup> | Hungary | RCT | Healthy aging adults (42/12) | 59.8 | 100 μT; 15 min ×5/wk; 12 wk | Systolic/diastolic BP; BMI; arterial stiffness |
| Kwan et al. (2015)<sup>37</sup> | China | RCT | Diabetic foot ulcer (7/6) | 62.7 | 1200 μT; 12 Hz; 60 min; 3 wk | Capillary diameter and velocity; skin blood flow |
| Kim et al. (2020)<sup>38</sup> | USA | RCT | Metabolic syndrome (23/21) | 58.3 | 390 μT; 30 Hz; 16 min thrice daily; 12 wk | Systolic/diastolic BP; nitric oxide |
| Sun et al. (2016)<sup>17</sup> | China | RCT | Diabetes (n = 22) + healthy (n = 21), each randomised to PEMF or sham | 67.4 | 500 μT; 12 Hz; single 30-min session | Blood-flow velocity; vein diameter; microcirculation |
| Biermann et al. (2020)<sup>23</sup> | Germany | Non-RCT (within-subject) | Healthy volunteers (15, within-subject) | 25.0 | 150 μT; 30 Hz; 40 min; 3 wk | Skin microcirculation |
| Nishimura et al. (2011)<sup>19</sup> | Japan | RCT | Mild-moderate hypertension (10/9) | 53.9 | 1 μT; 7 Hz; 12.5 min; 4 wk | Systolic/diastolic BP |
| Ahmad et al. (2021)<sup>26</sup> | Egypt | Non-RCT (interventional) | Peripheral artery disease (15/15) | 55.9 | 2000 μT; 15 Hz; 60 min; 8 wk | Ankle-brachial index |

*BMI, body mass index; BP, blood pressure; CRP, C-reactive protein; PEMF, pulsed electromagnetic field; RCT, randomised controlled trial. Superscript numbers refer to the reference list of the main manuscript. Full risk-of-bias and GRADE assessments are provided in the Supplementary Material (Tables S4 and S5).*

### Overall effect of PEMF

In our primary analysis, which pooled 23 outcomes from eight studies, PEMF was associated with a statistically significant overall benefit: the pooled effect favoured the intervention by 0.54 MCID units (95% CI 0.08 to 1.00). On this scale, zero denotes no effect and 1.0 one full clinically important difference, so the pooled estimate corresponds to roughly half of one clinically important difference, and its confidence interval excluded the null. Between-study heterogeneity was low to moderate (I^2^ = 36%; p = 0.144), suggesting a broadly consistent effect across studies (Figure 2).

**Figure 2.**
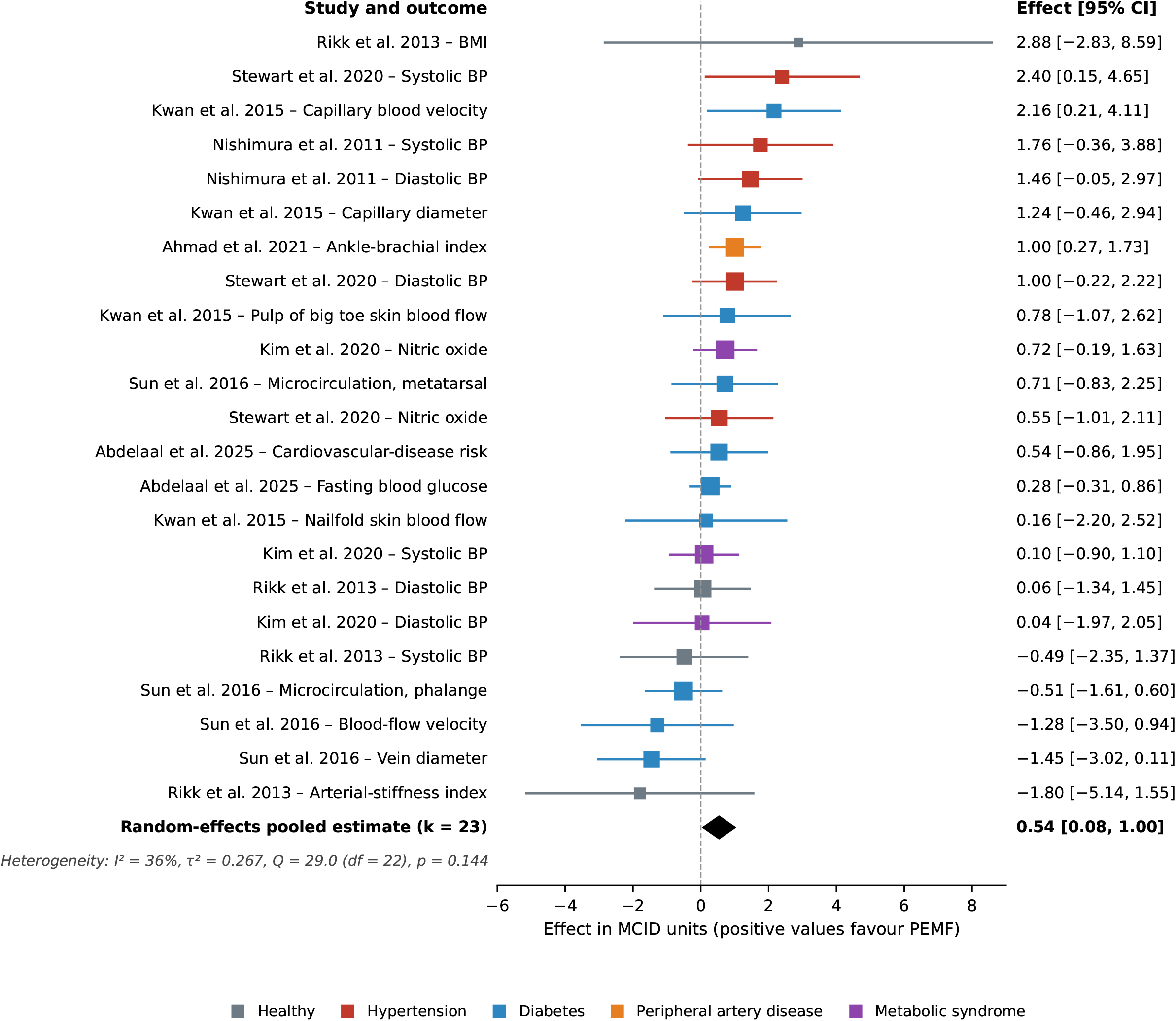
Primary forest plot of PEMF effects on cardiometabolic outcomes. Effects are expressed in MCID units (favourable direction positive), with the multilevel random-effects pooled estimate (k = 23 outcomes from eight studies) shown at the base. Squares are individual outcome estimates coloured by condition, with horizontal lines showing 95% confidence intervals; the diamond is the pooled estimate. BP, blood pressure; CI, confidence interval; MCID, minimal clinically important difference.

The clearest individual signals came from the three outcomes whose confidence intervals excluded the null, each a clear beneficial effect: systolic blood pressure (Stewart et al. 2020^35^), ankle-brachial index (Ahmad et al. 2021^26^), and capillary blood velocity (Kwan et al. 2015^37^), spanning blood pressure, peripheral arterial function, and microvascular flow. Of the remaining 20 outcomes, 15 had point estimates in the favourable direction and five in the unfavourable direction, none statistically significant; we present the complete outcome-level estimates in Figure 2 and Table S3.

### Blood pressure and hypertension

Our subgroup analysis suggested a particularly strong and consistent benefit in hypertension (Figure 3). Across five outcomes from double-blind trials, PEMF was associated with a large, statistically significant effect of 1.24 MCID units (95% CI 0.52 to 1.96), with no detectable heterogeneity (I^2^ = 0%; p = 0.692), representing more than one full clinically important improvement and an apparently reproducible signal. In diabetes (ten outcomes, three studies), the effect was smaller and did not reach significance (0.28 MCID units, 95% CI −0.61 to 1.17), and the between-subgroup difference was not statistically significant (p = 0.131). In the single metabolic syndrome trial (Kim et al. 2020^38^), the effects were close to null for systolic and diastolic blood pressure (0.10 and 0.04 MCID units) and non-significant for nitric oxide (0.72, −0.19 to 1.63; Table S3).

**Figure 3.**
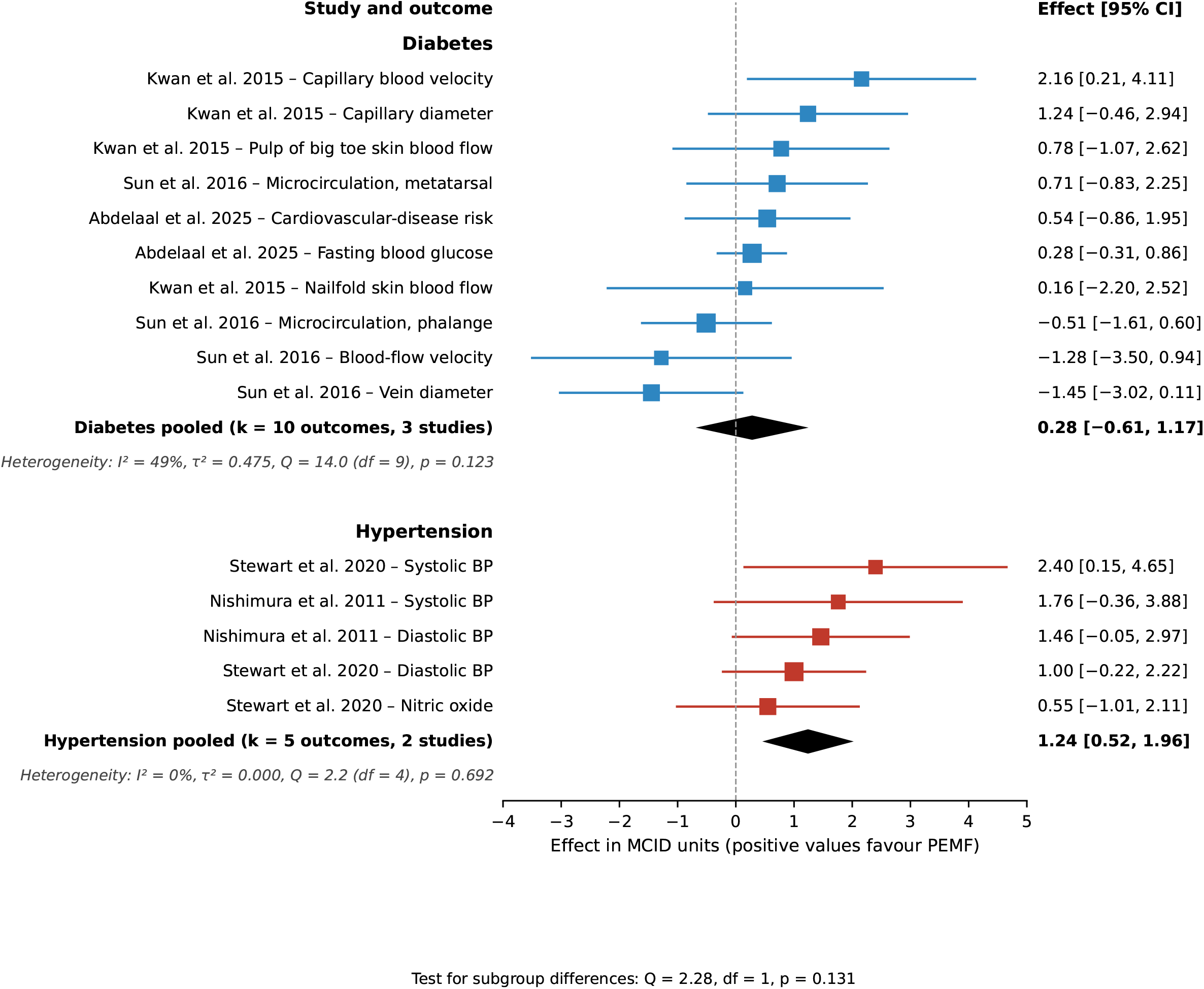
Subgroup forest plot (hypertension versus diabetes). Outcome-level estimates and pooled estimates for diabetes (10 outcomes, three studies) and hypertension (five outcomes, two studies), with per-subgroup heterogeneity and the test for subgroup differences. Effects are in MCID units, with positive values favouring PEMF.

### Sensitivity and dose-response analyses

In a sensitivity analysis restricted to the most rigorous trials, those with a double-blind, sham-controlled design delivering a multi-session treatment course (16 outcomes from five studies), the pooled effect was 0.74 MCID units (95% CI 0.20 to 1.28), statistically significant and with no heterogeneity (I^2^ = 0%) (Figure 4). A leave-one-out analysis of the primary model confirmed its stability: the pooled estimate remained consistent regardless of which study was removed (0.45 to 0.68 MCID units), with no single study exerting undue influence.

**Figure 4.**
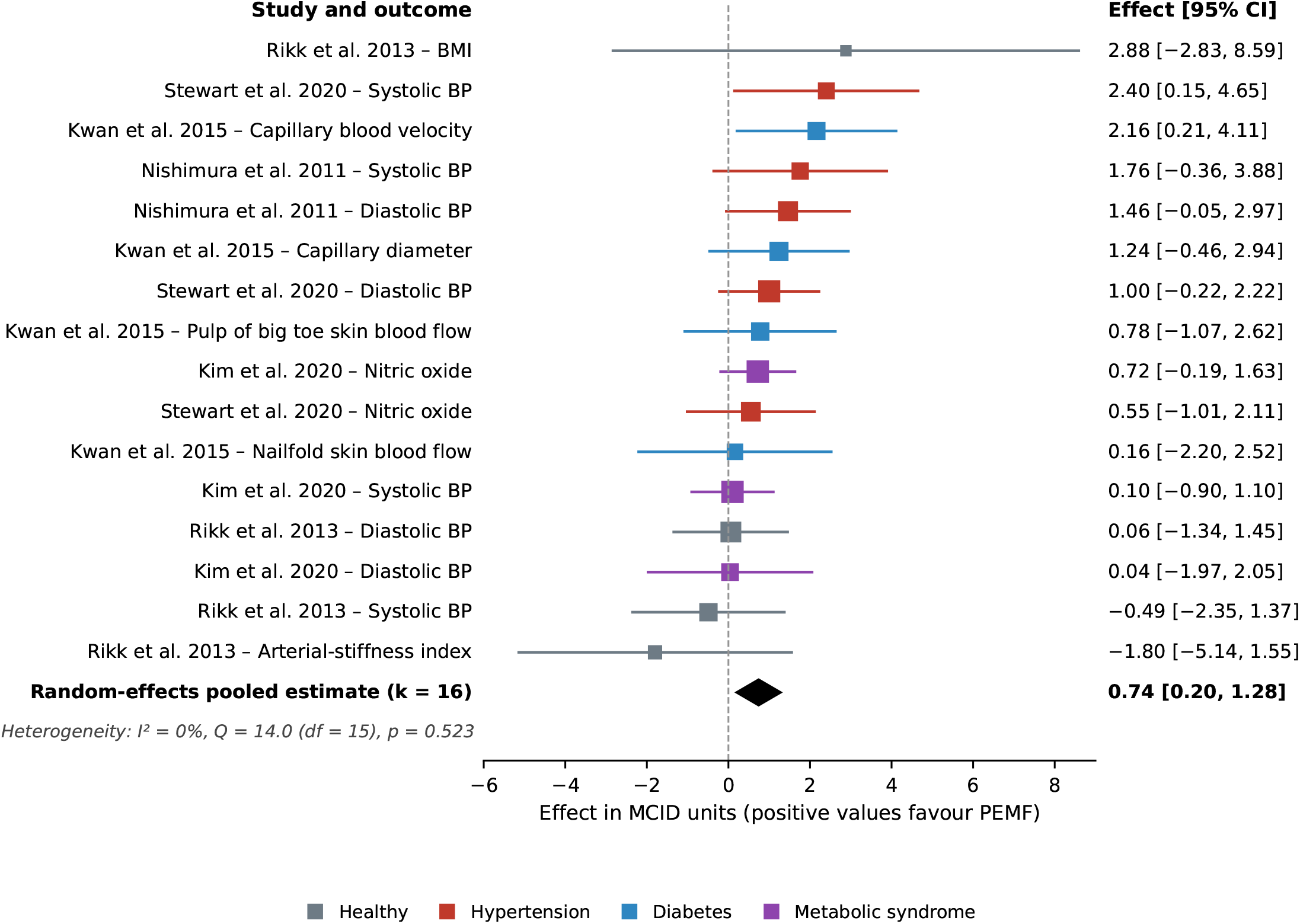
Sensitivity analysis restricted to the most rigorous trials. Pooled estimate restricted to double-blind, sham-controlled trials delivering a multi-session treatment course (16 outcomes from five studies); the effect strengthened to 0.74 MCID units (95% CI 0.20 to 1.28) with no heterogeneity (I^2^ = 0%). Squares are individual outcome estimates coloured by condition, with horizontal lines showing 95% confidence intervals; the diamond is the random-effects pooled estimate.

In univariable meta-regression, none of intensity, frequency, session duration, total exposure, or intervention duration was individually associated with effect size (all p > 0.05). In a multivariable model, however, two candidate moderators emerged: greater total exposure was associated with a larger treatment effect (β = 2.55; p = 0.028), whereas higher stimulation frequency was inversely associated with effect (β = −2.73; p = 0.026), with intensity non-significant after adjustment. Because these associations emerged only after mutual adjustment in a small number of studies, they were rated very low certainty and warrant confirmation in larger studies (univariable fits in Figures S1 to S3).

### Risk of bias

Of the eight randomised trials, two were judged at low overall risk of bias, five raised some concerns, and one (Wróbel et al. 2010^18^) was at high risk, driven by missing outcome data; both non-randomised studies were at moderate risk of bias (Table S4).

Egger’s test showed no evidence of funnel-plot asymmetry (p = 0.871). With fewer than 10 contributing studies this test is underpowered^32^, so we cannot fully exclude small-study effects. Certainty of evidence was moderate for the hypertension subgroup (Table S5).

## DISCUSSION

### Summary of key findings

In this systematic review and meta-analysis, PEMF was associated with a statistically significant improvement in cardiometabolic outcomes overall (about half a clinically important difference), most pronounced in blood pressure. The overall estimate was robust to restriction to double-blind, multi-session trials and to removal of any single study. Taken together, our findings suggest that PEMF may be a promising home-based non-pharmacological adjunct in cardiometabolic care, with its clearest and strongest role in blood-pressure lowering.

### Discussion of key findings

The overall benefit of PEMF in cardiometabolic diseases was expected as the proposed mechanisms have been suggested to modulate cellular activity, including transmembrane ion transport^9,10^, nitric-oxide signalling^9^, inflammatory pathways^11^, and microvascular perfusion^12^, the same processes disrupted in cardiometabolic disease^2^. The concentration of benefit in blood pressure is consistent with PEMF’s proposed mechanisms. The low-amplitude currents induced by PEMF are thought to act on the vascular endothelium, promoting nitric-oxide release^9^ and improving the reactivity of resistance arteries^12,35^, both principal determinants of blood pressure^7,8^. Because blood-pressure regulation depends heavily on this moment-to-moment vascular tone, a stimulus that enhances endothelial responsiveness would be expected to lower blood pressure more readily than it could reverse the more established metabolic and structural derangements of diabetes. The intermittent, exposure-recovery character of PEMF may be relevant here, since such patterns are typical of hormetic stimuli that elicit adaptive rather than maladaptive vascular remodelling^10,15^.

These findings extend a literature that has until now been fragmented across indications and outcomes. PEMF has an established evidence base in fracture healing^14^ and a growing one in osteoarthritis^8^ and wound care^8,37^, but its cardiometabolic effects had not been quantitatively synthesised. Individual trials and a systematic review had suggested a blood-pressure benefit^13,19,35^, yet the absence of a common scale prevented pooling; by expressing every outcome in MCID units we were able to show that the blood-pressure signal is not only present but the most consistent effect across the available evidence, and that a smaller but significant benefit persists when all cardiometabolic outcomes are combined.

The weaker effect in diabetes may reflect the structural remodelling of the diabetic vasculature^2,39^, which can blunt the very microvascular responsiveness that PEMF appears to engage, together with the limited power of a few small studies^30^. This pattern suggests the benefit may be greatest where vascular tone remains most modifiable, and supports evaluating PEMF condition by condition, beginning with hypertension.

Our meta-regression identified two candidate moderators of response: total exposure and stimulation frequency. Greater cumulative exposure was associated with larger effects, consistent with a dose-response relationship and with in vitro evidence that longer exposures more reliably elicit measurable cellular responses^7^. Within the 7 to 30 Hz range represented in the meta-analysis, lower frequencies were associated with greater clinical benefit, contrasting with the stronger cellular responses often reported at higher frequencies in vitro^7^; because the pooled trials employed comparatively low frequencies, the optimal window may differ between cellular and clinical settings. Because both associations appeared only after mutual adjustment, may partly reflect collinearity between exposure and frequency across few studies, and were rated very low certainty, they should be regarded as dosing hypotheses rather than findings, namely that longer cumulative exposure at lower frequencies may confer greater benefit, and they warrant direct testing in trials that compare frequencies and exposure durations.

### Clinical and public health relevance

The magnitude of the blood-pressure effect is potentially clinically relevant: the hypertension point estimate exceeded one clinically important difference, although its confidence interval (0.52 to 1.96) was compatible with smaller effects and the estimate combined blood-pressure and nitric-oxide outcomes. For systolic blood pressure specifically, the two contributing trials observed between-group differences of approximately 9 and 12 mmHg^19,35^, a magnitude comparable to single-agent antihypertensive therapy^40^, and reductions of this order are associated with meaningfully lower rates of stroke and myocardial infarction^22^. Because hypertension is the leading modifiable cause of cardiovascular mortality worldwide and control rates remain limited by poor adherence, a well-tolerated, non-invasive, self-administered (home-based) adjunct could carry public-health value^4–6^.

### Strengths and limitations

The principal strength of this synthesis is the use of MCID-standardised effect sizes, which allowed clinically heterogeneous endpoints to be pooled on a common, clinically interpretable scale. Unlike the conventional standardised mean difference, which rescales each effect by its own study’s standard deviation and therefore yields a unit with no fixed clinical meaning^21^, MCID standardisation holds the denominator constant across studies and renders the pooled estimate interpretable as multiples of a clinically important difference. The review was prospectively registered, and analytic decisions, including those not prespecified in the protocol, are documented in the Methods and Supplementary Material.

Several limitations should be acknowledged. The evidence base, although sufficient to detect a significant overall effect and a clear blood-pressure signal, remains small, and further trials would refine the precision of the estimates; reassuringly, the pooled effect remained stable in leave-one-out testing (0.45 to 0.68 MCID units) and strengthened when the analysis was restricted to the most rigorous double-blind, multi-session trials, indicating that the signal reflects the evidence as a whole rather than any single study. Where no published anchor existed, we assigned a distribution-based MCID of half the pooled within-arm standard deviation; for these outcomes the denominator is study-specific, so the effect is equivalent to twice the standardised mean difference and lacks the fixed clinical anchor of the remaining outcomes. The thresholds for fasting glucose, BMI, and the ankle-brachial index were set empirically rather than taken from published anchors, so effects on these outcomes should be interpreted with corresponding caution. The use of MCIDs was also not prespecified in the registered protocol. Effects were derived from end-of-intervention means, which in small trials may be influenced by baseline imbalance^30^. Two trials could not enter the quantitative synthesis because they did not report outcome-level dispersion, although they were retained in the qualitative synthesis, and adverse events were not systematically reported across trials, so conclusions on tolerability remain provisional. The two contributing trials of Stewart et al. 2020^35^ and Kim et al. 2020^38^ were conducted by the same research group with the same device and protocol, so some overlap of participants cannot be excluded. Finally, as is typical of an emerging field, the small number of studies limited formal testing for publication bias^32^. We therefore interpret these results as a foundation for confirmatory work rather than a definitive synthesis.

## CONCLUSION

PEMF therapy was associated with a statistically significant improvement in cardiometabolic outcomes overall, of about half a clinically important difference, most pronounced in blood pressure. Our findings suggest that PEMF may be a potential, home-based, non-invasive adjunct in cardiometabolic care, with its clearest role in blood-pressure lowering in hypertension.

## DECLARATIONS

### Ethics approval

This study did not involve data collection; ethics approval and informed consent were therefore not required.

### Author contributions

F.P.C. and R.R.S. conceived and designed the study, conducted the literature search, screening, and data extraction, performed the statistical analysis, and wrote the manuscript. P.A. served as the third reviewer, adjudicating disagreements during study selection and extraction. All authors reviewed the content and approved the final manuscript.

### Data availability

The study did not generate any data but rather collated available published studies which have been referenced.

### Registration

The protocol was prospectively registered in PROSPERO (CRD420261404575).

