## Supplementary Appendix for "Home-Based Pulsed Electromagnetic Field Therapy for Blood Pressure and Cardiometabolic Health: A Systematic Review and Meta-Analysis"

### **Contents**

**Table S1.** Search strategy for each database

**Table S2.** Detailed characteristics of the included studies

**Table S3.** Full outcome-level effect estimates

**Figure S1.** Meta-regression bubble plot: total exposure

**Figure S2.** Meta-regression bubble plot: stimulation frequency

**Figure S3.** Meta-regression bubble plot: field intensity

**Table S4.** Risk-of-bias assessment (RoB 2 and ROBINS-I)

**Table S5.** GRADE summary of findings

*All estimates are expressed in units of the minimal clinically important difference (MCID), with a positive value indicating a clinically favourable effect. The complete analytic dataset and R analysis script are available from the authors on request.*

***Table S1. Search strategy for each database.***

| **Database** | **Search strategy** |
| --- | --- |
| PubMed (MEDLINE) | ("pulsed electromagnetic field*"[tiab] OR "pulsed magnetic field*"[tiab] OR "pulsed electromagnetic therap*"[tiab] OR PEMF[tiab]) AND (hypertens*[tiab] OR "blood pressure"[tiab] OR diabet*[tiab] OR glucose[tiab] OR HbA1c[tiab] OR insulin[tiab] OR obes*[tiab] OR "metabolic syndrome"[tiab] OR cardiometabolic[tiab] OR vascular[tiab] OR endothelial[tiab]) AND Humans[Mesh] |
| Embase | ('pulsed electromagnetic field*':ti,ab OR 'pulsed magnetic field*':ti,ab OR 'pulsed electromagnetic therap*':ti,ab OR PEMF:ti,ab) AND ('hypertension'/exp OR 'diabetes mellitus'/exp OR 'metabolic syndrome'/exp OR 'obesity'/exp OR hypertens*:ti,ab OR 'blood pressure':ti,ab OR diabet*:ti,ab OR glucose:ti,ab OR insulin:ti,ab OR obes*:ti,ab OR cardiometabolic:ti,ab OR vascular:ti,ab OR endothelial:ti,ab) AND [humans]/lim |
| Web of Science | TS=(("pulsed electromagnetic field*" OR "pulsed magnetic field*" OR "pulsed electromagnetic therap*" OR PEMF) AND (hypertension OR "blood pressure" OR diabetes OR glucose OR insulin OR obesity OR "metabolic syndrome" OR cardiometabolic OR vascular OR endothelial)) |
| Scopus | TITLE-ABS-KEY(("pulsed electromagnetic field*" OR "pulsed magnetic field*" OR "pulsed electromagnetic therap*" OR PEMF) AND (hypertension OR "blood pressure" OR diabetes OR glucose OR insulin OR obesity OR "metabolic syndrome" OR cardiometabolic OR vascular OR endothelial)) |
| Cochrane CENTRAL | ("pulsed electromagnetic field*" OR "pulsed magnetic field*" OR "pulsed electromagnetic therap*" OR PEMF) AND (hypertension OR "blood pressure" OR diabetes OR glucose OR insulin OR obesity OR "metabolic syndrome" OR cardiometabolic OR vascular OR endothelial) |
| ClinicalTrials.gov | Condition or disease: Hypertension OR Diabetes OR Metabolic Syndrome OR Obesity OR Cardiovascular Disease. Other terms: Pulsed electromagnetic OR PEMF. Filters: Completed studies; adults 18 years and older. |
| WHO ICTRP | Pulsed electromagnetic AND hypertension; Pulsed electromagnetic AND diabetes; PEMF AND metabolic. Filters: adult studies. |

*Searches were conducted without restrictions on publication year, and human filters were applied where available. Terms combined controlled vocabulary (MeSH and Emtree) with free-text terms for the intervention and for cardiometabolic conditions, adapted to each database’s syntax. Google Scholar and the reference lists of included studies were searched separately.*

***Table S2. Detailed characteristics of the included studies.***

| **Study (year)** | **Country** | **Design** | **Condition (n PEMF/control)** | **Age (y)** | **Intensity (µT)** | **Freq (Hz)** | **Session (min)** | **Weeks** | **Total (min)** | **Key outcomes** |
| --- | --- | --- | --- | --- | --- | --- | --- | --- | --- | --- |
| Abdelaal et al. (2025)^34^ | Saudi Arabia | RCT | Type 2 diabetes (14/14) | 57.8 | 2000 | 15 | 30 | 8 | 1680 | Fasting glucose; CVD risk |
| Wróbel et al. (2010)^18^ | Poland | RCT | Diabetic polyneuropathy (32/29) + healthy (20, PEMF only) | 49.8 | 100 | 188.5 | 20 | 3 | 300 | Defensin; CRP |
| Stewart et al. (2020)^35^ | USA | RCT | Hypertension (15/11) | 59.5 | 390 | 30 | 16 | 12 | 4032 | Systolic/diastolic BP; nitric oxide |
| Rikk et al. (2013)^36^ | Hungary | RCT | Healthy aging adults (42/12) | 59.8 | 100 | NR | 15 | 12 | 900 | Systolic/diastolic BP; BMI; arterial stiffness |
| Kwan et al. (2015)^37^ | China | RCT | Diabetic foot ulcer (7/6) | 62.7 | 1200 | 12 | 60 | 3 | 900 | Capillary diameter and velocity; skin blood flow |
| Kim et al. (2020)^38^ | USA | RCT | Metabolic syndrome (23/21) | 58.3 | 390 | 30 | 16 | 12 | 4032 | Systolic/diastolic BP; nitric oxide |
| Sun et al. (2016)^17^ | China | RCT | Diabetes (n = 22) + healthy (n = 21), each randomised to PEMF or sham | 67.4 | 500 | 12 | 30 | single | 30 | Blood-flow velocity; vein diameter; microcirculation |
| Biermann et al. (2020)^23^ | Germany | Non-RCT (within-subject) | Healthy volunteers (15) | 25.0 | 150 | 30 | 40 | 3 | 420 | Skin microcirculation |
| Nishimura et al. (2011)^19^ | Japan | RCT | Mild-moderate hypertension (10/9) | 53.9 | 1 | 7 | 12.5 | 4 | 125 | Systolic/diastolic BP |
| Ahmad et al. (2021)^26^ | Egypt | Non-RCT (interventional) | Peripheral artery disease (15/15) | 55.9 | 2000 | 15 | 60 | 8 | 1440 | Ankle-brachial index |

*BMI, body mass index; BP, blood pressure; CRP, C-reactive protein; CVD, cardiovascular disease; NR, not reported; PEMF, pulsed electromagnetic field; RCT, randomised controlled trial. Total exposure = single-session minutes × sessions per week × weeks. Superscript numbers refer to the reference list of the main manuscript.*

***Table S3. Full outcome-level MCID-standardised effect estimates (favourable direction positive), ordered by effect size.***

| **Study (year)** | **Outcome** | **Condition** | **Effect (MCID units) (95% CI)** |
| --- | --- | --- | --- |
| Rikk et al. (2013) | BMI | Healthy | 2.88 (−2.83 to 8.59) |
| Stewart et al. (2020) | Systolic BP | Hypertension | 2.40 (0.15 to 4.65) |
| Kwan et al. (2015) | Capillary blood velocity | Diabetes | 2.16 (0.21 to 4.11) |
| Nishimura et al. (2011) | Systolic BP | Hypertension | 1.76 (−0.36 to 3.88) |
| Nishimura et al. (2011) | Diastolic BP | Hypertension | 1.46 (−0.05 to 2.97) |
| Kwan et al. (2015) | Capillary diameter | Diabetes | 1.24 (−0.46 to 2.94) |
| Ahmad et al. (2021) | Ankle-brachial index | Peripheral artery disease | 1.00 (0.27 to 1.73) |
| Stewart et al. (2020) | Diastolic BP | Hypertension | 1.00 (−0.22 to 2.22) |
| Kwan et al. (2015) | Pulp of big toe skin blood flow | Diabetes | 0.78 (−1.07 to 2.62) |
| Kim et al. (2020) | Nitric oxide | Metabolic syndrome | 0.72 (−0.19 to 1.63) |
| Sun et al. (2016) | Microcirculation, metatarsal | Diabetes | 0.71 (−0.83 to 2.25) |
| Stewart et al. (2020) | Nitric oxide | Hypertension | 0.55 (−1.01 to 2.11) |
| Abdelaal et al. (2025) | Cardiovascular-disease risk | Diabetes | 0.54 (−0.86 to 1.95) |
| Abdelaal et al. (2025) | Fasting blood glucose | Diabetes | 0.28 (−0.31 to 0.86) |
| Kwan et al. (2015) | Nailfold skin blood flow | Diabetes | 0.16 (−2.20 to 2.52) |
| Kim et al. (2020) | Systolic BP | Metabolic syndrome | 0.10 (−0.90 to 1.10) |
| Rikk et al. (2013) | Diastolic BP | Healthy | 0.06 (−1.34 to 1.45) |
| Kim et al. (2020) | Diastolic BP | Metabolic syndrome | 0.04 (−1.97 to 2.05) |
| Rikk et al. (2013) | Systolic BP | Healthy | −0.49 (−2.35 to 1.37) |
| Sun et al. (2016) | Microcirculation, phalange | Diabetes | −0.51 (−1.61 to 0.60) |
| Sun et al. (2016) | Blood-flow velocity | Diabetes | −1.28 (−3.50 to 0.94) |
| Sun et al. (2016) | Diameter of vein | Diabetes | −1.45 (−3.02 to 0.11) |
| Rikk et al. (2013) | Arterial-stiffness index | Healthy | −1.80 (−5.14 to 1.55) |

| **Pooled estimate** |  |  | **Effect (MCID units) (95% CI)** |
| --- | --- | --- | --- |
| Overall (k = 23) |  |  | 0.54 (0.08 to 1.00) |
| Hypertension subgroup (k = 5) |  |  | 1.24 (0.52 to 1.96) |
| Diabetes subgroup (k = 10) |  |  | 0.28 (−0.61 to 1.17) |

*Positive values favour PEMF. Confidence intervals excluding zero indicate a statistically detectable effect.*

**Figure S1. Meta-regression bubble plot: total exposure.** Exploratory relationship between cumulative PEMF exposure (minutes) and MCID-standardised effect size; bubble size is proportional to outcome weight (1/SE); the line shows the univariable meta-regression fit with its 95% confidence band.

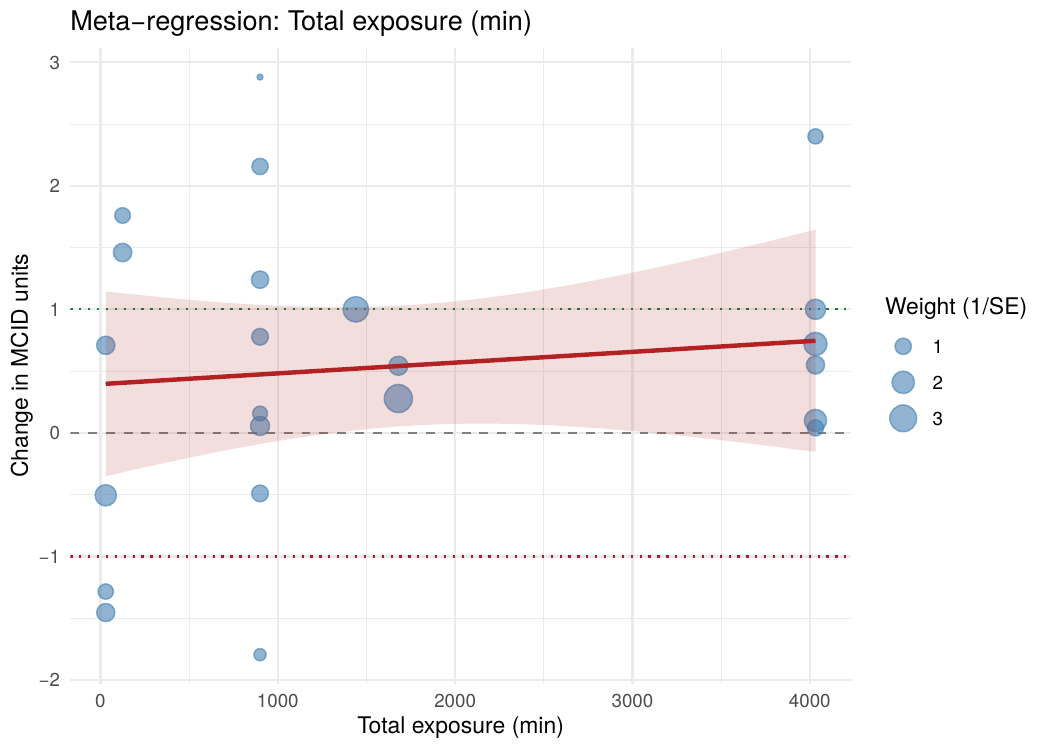

**Figure S2. Meta-regression bubble plot: stimulation frequency.** Exploratory relationship between PEMF frequency (Hz) and MCID-standardised effect size; bubble size and fitted line as in Figure S1. Rikk et al. (2013), which did not report frequency, is not shown.

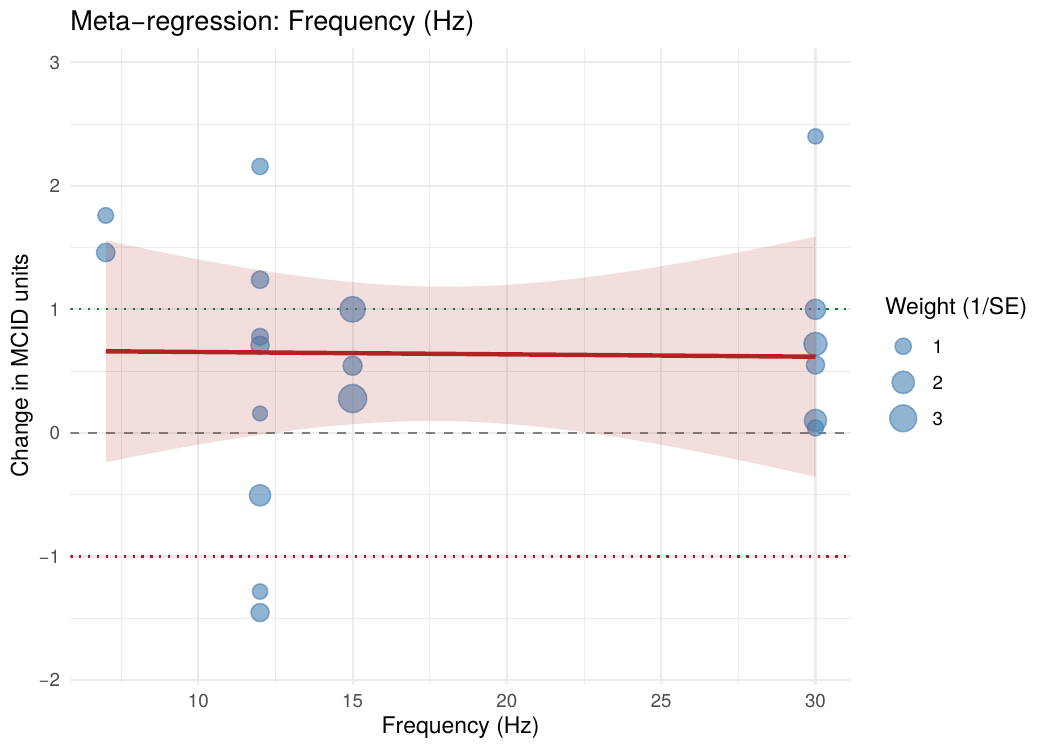

**Figure S3. Meta-regression bubble plot: field intensity.** Exploratory relationship between PEMF intensity (µT) and MCID-standardised effect size; bubble size and fitted line as in Figure S1.

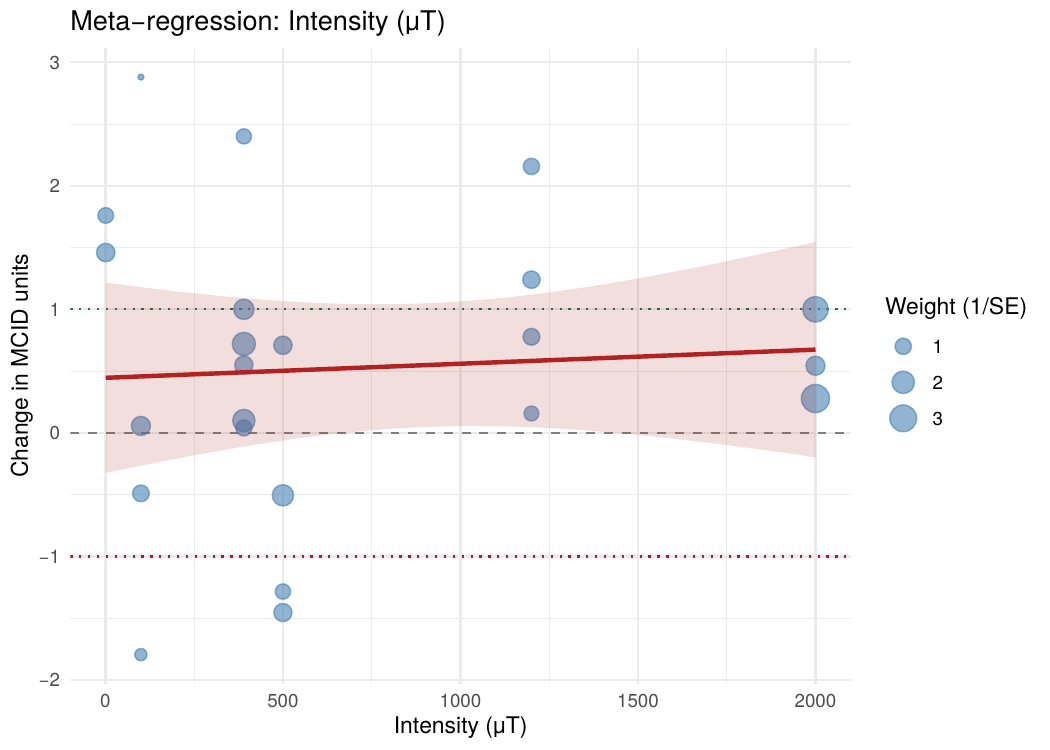

***Table S4. Risk-of-bias assessment. Randomised trials were assessed with Cochrane RoB 2; non-randomised studies with ROBINS-I.***

| **Study (year)** | **Design / tool** | **D1** | **D2** | **D3** | **D4** | **D5** | **Overall** |
| --- | --- | --- | --- | --- | --- | --- | --- |
| Abdelaal et al. (2025) | RCT (RoB 2) | Low | Some concerns | Low | Low | Low | Some concerns |
| Wróbel et al. (2010) | RCT (RoB 2) | Low | Low | High | Low | Some concerns | High |
| Stewart et al. (2020) | RCT (RoB 2) | Low | Low | Some concerns | Low | Low | Some concerns |
| Rikk et al. (2013) | RCT (RoB 2) | Some concerns | Low | Low | Low | Some concerns | Some concerns |
| Kwan et al. (2015) | RCT (RoB 2) | Low | Low | Low | Low | Some concerns | Some concerns |
| Kim et al. (2020) | RCT (RoB 2) | Low | Low | Low | Low | Low | Low |
| Sun et al. (2016) | RCT (RoB 2) | Low | Low | Low | Low | Some concerns | Some concerns |
| Biermann et al. (2020) | Non-RCT (ROBINS-I) | Moderate | Low | Low | Low | Moderate | Moderate |
| Nishimura et al. (2011) | RCT (RoB 2) | Low | Low | Low | Low | Low | Low |
| Ahmad et al. (2021) | Non-RCT (ROBINS-I) | Moderate | Low | Low | Moderate | Moderate | Moderate |

*RoB 2 domains: D1, randomisation process; D2, deviations from intended interventions; D3, missing outcome data; D4, measurement of the outcome; D5, selection of the reported result. ROBINS-I has seven domains, which were condensed into the five columns as follows: D1, bias due to confounding and selection of participants; D2, bias in classification of interventions and deviations from intended interventions; D3, missing data; D4, measurement of outcomes; D5, selection of the reported result. Two studies (Wróbel et al. 2010; Biermann et al. 2020) did not report dispersion data and contributed to the qualitative synthesis only.*

***Table S5. GRADE summary of findings for the overall estimate, subgroups, and dose-response meta-regression.***

| **Outcome / analysis** | **Effect (95% CI)** | **Risk of bias** | **Inconsistency** | **Indirectness** | **Imprecision** | **Publication bias** | **Certainty** |
| --- | --- | --- | --- | --- | --- | --- | --- |
| Overall pooled effect (8 studies, 23 outcomes) | 0.54 (0.08 to 1.00) | Some concerns to Low | Not serious (I² = 36%) | Not serious | Serious | Suspected (k < 10) | Low |
| Hypertension subgroup (2 studies, 5 outcomes) | 1.24 (0.52 to 1.96) | Some concerns to Low | Not serious | Not serious | Serious (few studies) | Unknown (k = 2) | Moderate |
| Diabetes subgroup (3 studies, 10 outcomes) | 0.28 (−0.61 to 1.17) | Some concerns | Some concerns (not downgraded) | Some concerns (not downgraded) | Serious (CI spans zero) | Unknown | Low |
| Dose-response meta-regression | Exploratory | As above | N/A | N/A | Very serious (k < 10 per predictor) | N/A | Very low |

*GRADE certainty reflects the strength of evidence, not the direction of effect; all pooled point estimates are in the favourable direction. The hypertension subgroup carries the highest certainty (Moderate) and represents the most promising clinical signal warranting confirmatory trials.*
